# Determinants of Skilled Birth Attendance in Nigeria: A Population-Based Analysis of the 2018 Demographic and Health Survey

**DOI:** 10.64898/2026.04.23.26350432

**Authors:** Ugoeze Lucy Unegbu

## Abstract

**Background:** Nigeria accounts for approximately 19% of global maternal deaths, yet skilled birth attendance (SBA) coverage stood at only 44.9% in 2018. Understanding the independent determinants of SBA after controlling for confounding is essential for evidence-based policy prioritisation.

**Methods:** We conducted a cross-sectional analysis of 21,465 women with a birth in the five years preceding the 2018 Nigeria Demographic and Health Survey (NDHS). Survey-weighted multivariable logistic regression was used to estimate adjusted odds ratios (aOR) for seven sociodemographic predictors of SBA. Confounding was quantified by comparing crude and adjusted estimates.

**Results:** Overall SBA prevalence was 44.9%, ranging from 17.7% in the North West to 85.6% in the South West. Higher education (aOR = 7.01, 95% CI: 5.68–8.67), richest wealth quintile (aOR = 6.27, 95% CI: 5.27–7.46), and attending ≥4 antenatal care (ANC) visits (aOR = 3.80, 95% CI: 3.51–4.11) were the strongest independent predictors. Confounding was substantial: 89.0% of education’s crude effect and 87.1% of the wealth effect were attributable to correlated socioeconomic factors. ANC utilisation showed the least confounding (56.3% attenuation), consistent with a more direct causal pathway.

**Conclusions:** ANC utilisation is the most modifiable and directly actionable determinant of skilled birth attendance in Nigeria. Geographically targeted investment in ANC coverage, demand-side financing, and girls’ education are urgently needed to close Nigeria’s SBA gap.

## 1. Introduction

Maternal mortality remains a defining global health challenge, with the vast majority of deaths concentrated in low- and middle-income countries (LMICs). Nigeria is among the highest-burden settings worldwide, with an estimated 512 maternal deaths per 100,000 live births in 2017, accounting for approximately 19% of all global maternal deaths (World Health Organization [WHO], 2019). Most of these deaths are attributable to direct obstetric causes such as hemorrhage, sepsis, hypertensive disorders, and obstructed labor, complications that are largely preventable with skilled care at the time of delivery (Say et al., 2014). Despite global progress in maternal health, Nigeria continues to lag behind, and the country is unlikely to meet the Sustainable Development Goal target of reducing maternal mortality to fewer than 70 per 100,000 live births by 2030 (United Nations, 2015) without substantial improvements in the coverage and quality of skilled delivery care.

Skilled birth attendance is defined as delivery by a doctor, nurse, midwife, or other trained health professional. It is a cornerstone intervention for reducing maternal and neonatal mortality (WHO, 2004). Yet, in Nigeria, national SBA coverage stood at approximately 44.9% in 2018, masking extreme geographic and socioeconomic heterogeneity (National Population Commission [NPC] & ICF, 2019). In the North West geopolitical zone, fewer than one in five women deliver with skilled care, while in parts of the South West, coverage exceeds 85%. This internal disparity signals that population-level averages obscure the depth of inequity and the urgency for regionally differentiated policy responses.

A substantial body of evidence has identified education, household wealth, rural residence, parity, and ANC utilization as correlates of SBA in sub-Saharan African settings (Adewuyi et al., 2017; Moyer & Mustafa, 2013). Since education, wealth, and residence are highly correlated, crude estimates can be severely inflated and may misattribute the contribution of individual determinants. This limits their utility for policy prioritization. Understanding the independent contribution of each factor, after accounting for correlated exposures is critical for designing interventions that efficiently target the proximal drivers of low SBA uptake.

This study addresses this gap by conducting a multivariable analysis of nationally representative data from the 2018 NDHS, with explicit assessment of confounding magnitude. The objectives were:

1. to determine the prevalence of SBA in Nigeria in 2018;
2. to examine crude associations between sociodemographic factors and SBA;
3. to identify independent predictors of SBA after adjustment for confounders; and
4. to quantify the extent of confounding in key associations.

## 2. Methods

### 2.1 Study Design and Data Source

This was a cross-sectional analysis of secondary data from the 2018 Nigeria Demographic and Health Survey (NDHS). The NDHS is a nationally representative household survey conducted by the National Population Commission (NPC) with technical support from ICF under the DHS Program. The survey used a stratified two-stage cluster sampling design: in the first stage, enumeration areas (clusters) were selected with probability proportional to size; in the second stage, households were systematically sampled within each cluster. Women aged 15-49 years residing in selected households were eligible for the individual women’s questionnaire. Full methodological details are reported in the official survey report (NPC & ICF, 2019). DHS data are de-identified and publicly available upon registration at dhsprogram.com. No additional ethical approval was required for this secondary analysis, as the study used only openly available, de-identified data collected under the DHS program’s standard ethical procedures.

### 2.2 Study Population and Analytic Sample

The individual recode file (NGIR7BFL.DTA) contained records for 41,821 women aged 15-49. The analytic sample was restricted to women who reported a live birth in the five years preceding the survey (n = 21,792), consistent with the DHS approach of collecting birth history for the most recent birth within this window. Women with missing data on ANC visits (n = 327, 1.5%) were excluded, yielding a final analytic sample of 21,465 women.

### 2.3 Outcome Variable

The primary outcome was skilled birth attendance, defined as delivery assisted by a doctor, nurse/midwife, or auxiliary midwife (DHS variables m3a_1, m3b_1, m3c_1). SBA was coded as a binary variable: 1 if any skilled attendant type was present at delivery, 0 otherwise. Delivery attended only by a traditional birth attendant, relative, or no attendant was coded as no SBA.

### 2.4 Explanatory Variables

Seven explanatory variables were included based on a priori theoretical and empirical grounds:

- **Maternal education** (v106): categorized as no education, primary, secondary, and higher
- **Household wealth index** (v190): a composite asset-based index constructed by DHS, categorized into quintiles (poorest, poorer, middle, richer, richest)
- **Place of residence** (v025): urban or rural
- **Geopolitical region** (v024): six zones (North West, North East, North Central, South West, South East, South South)
- **Maternal age** (v012): grouped into six five-year categories (15-19, 20-24, 25-29, 30-34, 35-39, 40-49)
- **Parity** (v201): grouped as 1, 2-3, 4-5, and 6 or more children
- **ANC utilization** (m14_1): dichotomized as fewer than 4 visits versus 4 or more visits, consistent with the WHO minimum recommendation for focused antenatal care (WHO, 2016)

### 2.5 Statistical Analysis

Descriptive statistics were computed to characterize the sample distribution across all covariates. SBA prevalence was calculated overall and stratified by each explanatory variable. Chi-square tests were used to assess bivariate associations between each predictor and SBA, with Cramer’s V computed as a measure of association strength.

Logistic regression was used to estimate the association between each predictor and SBA. Unadjusted (crude) odds ratios were estimated from simple logistic regression models for each predictor individually. Adjusted odds ratios (aOR) were estimated from a single multivariable logistic model including all seven predictors simultaneously. All models were fitted with sampling weights applied (v005/1,000,000) to account for the DHS complex survey design and produce nationally representative estimates. Odds ratios were exponentiated from log-odds coefficients, and 95% confidence intervals were derived using profile likelihood. P-values were obtained from Wald z-tests, with a significance level of alpha = 0.05. Model fit was assessed using McFadden’s pseudo R-squared.

Confounding was assessed by calculating the percentage change between crude and adjusted odds ratios for each predictor: ((aOR - crude OR) / crude OR) x 100. Multicollinearity was assessed using variance inflation factors (VIF). All analyses were conducted in Python using the pandas, statsmodels, and scipy libraries.

## 3. Results

### 3.1 Sample Characteristics

The analytic sample comprised 21,465 women with a birth in the five years preceding the 2018 NDHS. The majority resided in rural areas (64.6%), and the largest education category was no education (43.7%). Wealth was approximately uniformly distributed across quintiles by design. The North West zone contributed the largest proportion of the sample (29.0%). The mean maternal age was 29.7 years (SD = 7.2), and the mean parity was 4.1 children (SD = 2.6). Full sample characteristics are presented in Table 1.

**Table 1:**
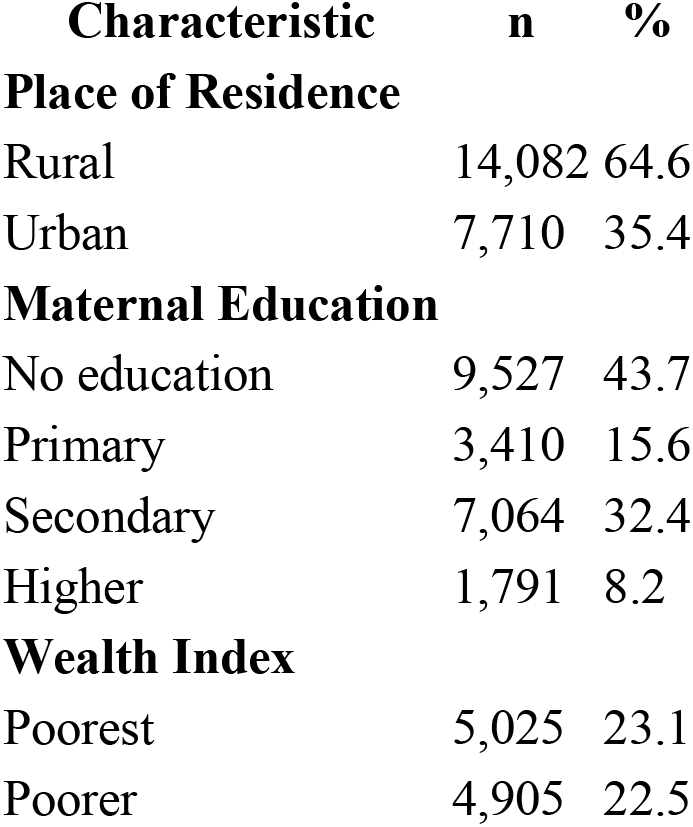
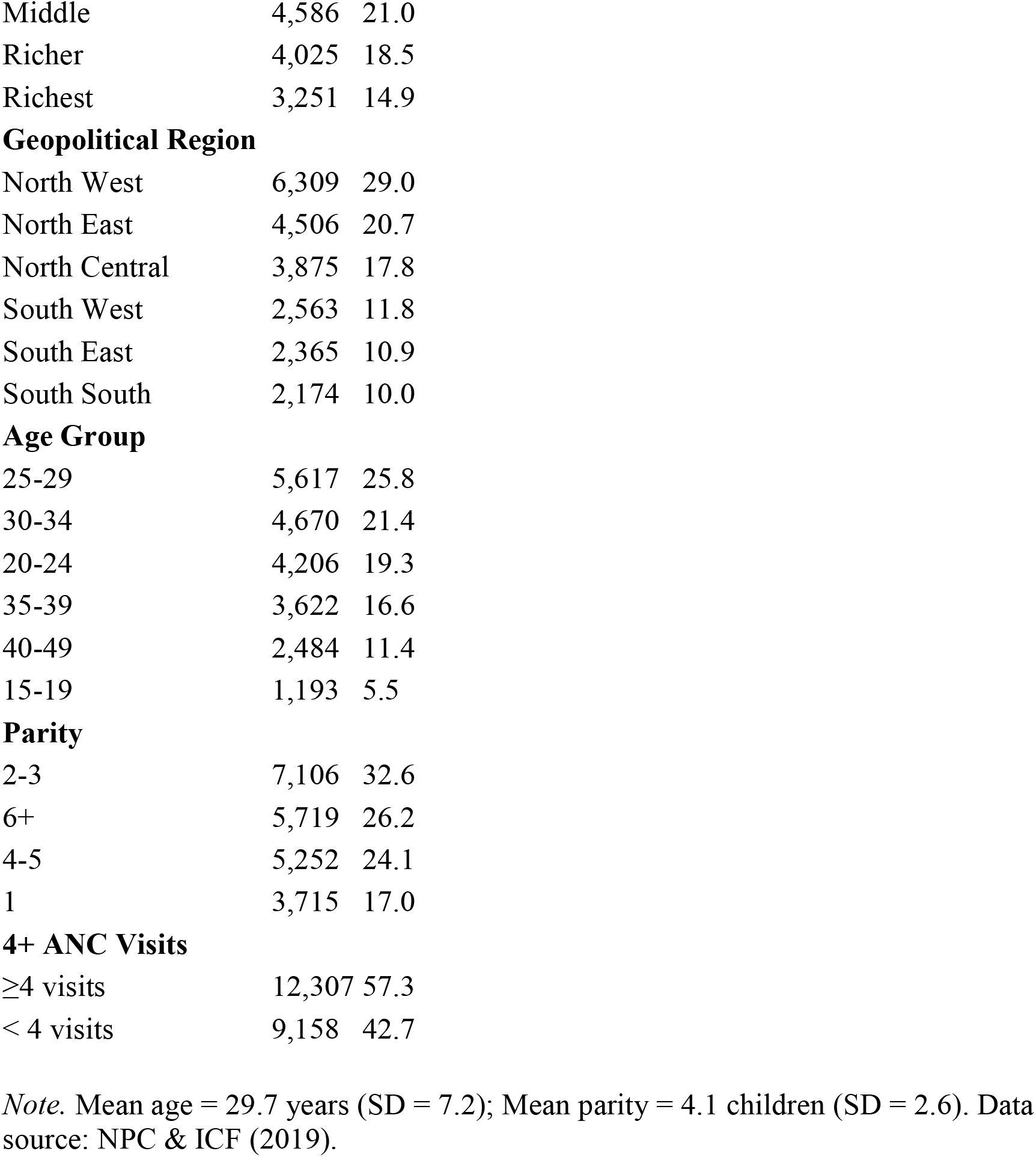
Sociodemographic characteristics of the analytic sample (n = 21,465)

| Characteristic | n | % |
| --- | --- | --- |
| **Place of Residence** |  |  |
| Rural | 14,082 | 64.6 |
| Urban | 7,710 | 35.4 |
| **Maternal Education** |  |  |
| No education | 9,527 | 43.7 |
| Primary | 3,410 | 15.6 |
| Secondary | 7,064 | 32.4 |
| Higher | 1,791 | 8.2 |
| **Wealth Index** |  |  |
| Poorest | 5,025 | 23.1 |
| Poorer | 4,905 | 22.5 |
| Middle | 4,586 | 21.0 |
| Richer | 4,025 | 18.5 |
| Richest | 3,251 | 14.9 |
| **Geopolitical Region** |  |  |
| North West | 6,309 | 29.0 |
| North East | 4,506 | 20.7 |
| North Central | 3,875 | 17.8 |
| South West | 2,563 | 11.8 |
| South East | 2,365 | 10.9 |
| South South | 2,174 | 10.0 |
| **Age Group** |  |  |
| 25-29 | 5,617 | 25.8 |
| 30-34 | 4,670 | 21.4 |
| 20-24 | 4,206 | 19.3 |
| 35-39 | 3,622 | 16.6 |
| 40-49 | 2,484 | 11.4 |
| 15-19 | 1,193 | 5.5 |
| **Parity** |  |  |
| 2-3 | 7,106 | 32.6 |
| 6+ | 5,719 | 26.2 |
| 4-5 | 5,252 | 24.1 |
| 1 | 3,715 | 17.0 |
| **4+ ANC Visits** |  |  |
| ≥4 visits | 12,307 | 57.3 |
| < 4 visits | 9,158 | 42.7 |

*Note.* Mean age = 29.7 years (SD = 7.2); Mean parity = 4.1 children (SD = 2.6). Data source: NPC & ICF (2019).

### 3.2 Prevalence of Skilled Birth Attendance

The overall weighted prevalence of SBA was 44.9% (n = 9,795). Marked variation was observed across all predictors (Table 2). SBA prevalence increased in a clear dose-response pattern with both education (15.7% for no education to 92.3% for higher education) and household wealth (12.2% in the poorest quintile to 87.2% in the richest quintile). Urban women had more than twice the SBA prevalence of rural women (68.4% vs. 32.1%). The regional disparity was striking: SBA ranged from 17.7% in the North West to 85.6% in the South West, a 68 percentage point gap within a single country. Women attending 4 or more ANC visits had substantially higher SBA prevalence than those attending fewer visits (64.6% vs. 17.4%). All associations were statistically significant at p < .001 (Table 2).

**Table 2:** SBA prevalence and bivariate associations with sociodemographic predictors.

| **Variable** | **SBA (%)** | **Chi-square (df)** | **p-value** |
| --- | --- | --- | --- |
| **Education** |  | chi-sq = 6835.30 (3) < .001 |  |
| No education | 15.7 |  |  |
| Primary | 48.1 |  |  |
| Secondary | 70.8 |  |  |
| Higher | 92.3 |  |  |
| **Wealth Index** |  | chi-sq = 6198.50 (4) < .001 |  |
| Poorest | 12.2 |  |  |
| Poorer | 26.2 |  |  |
| Middle | 49.7 |  |  |
| Richer | 69.0 |  |  |
| Richest | 87.2 |  |  |
| **Residence** |  | chi-sq = 2654.54 (1) < .001 |  |
| Rural | 32.1 |  |  |
| Urban | 68.4 |  |  |
| **ANC Visits** |  | chi-sq = 4746.69 (1) < .001 |  |
| < 4 visits | 17.4 |  |  |
| 4 or more visits | 64.6 |  |  |
| **Region** |  | chi-sq = 6129.76 (5) < .001 |  |
| North West | 17.7 |  |  |
| North East | 24.8 |  |  |
| North Central | 54.5 |  |  |
| South South | 58.0 |  |  |
| South East | 84.3 |  |  |
| South West | 85.6 |  |  |

### 3.3 Multivariable Logistic Regression

Results from the fully adjusted model are presented in Table 3. All predictors remained independently associated with SBA after mutual adjustment. Higher education was the strongest predictor overall (aOR = 7.01, 95% CI [5.68, 8.67], p < .001), followed by belonging to the richest wealth quintile (aOR = 6.27, 95% CI [5.27, 7.46], p < .001) and residing in the South East (aOR = 6.55, 95% CI [5.62, 7.63], p < .001) or South West (aOR = 6.37, 95% CI [5.46, 7.43], p < .001) relative to the North West.

**Table 3:**
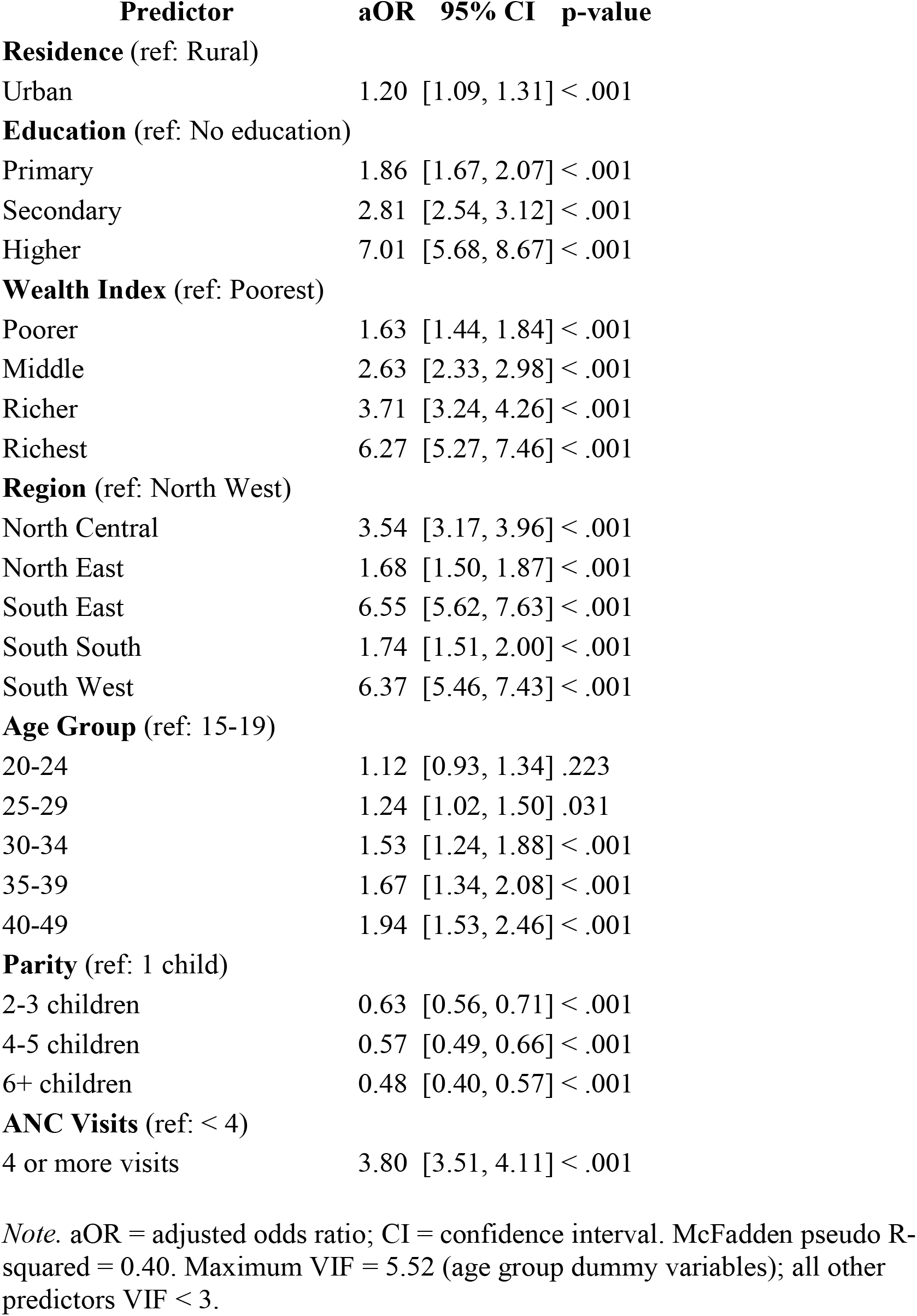
Multivariable logistic regression: adjusted odds ratios for skilled birth attendance (n = 21,465)

Attending 4 or more ANC visits was associated with nearly four times the odds of SBA (aOR = 3.80, 95% CI [3.51, 4.11], p < .001). After adjustment, urban residence had a substantially attenuated but still significant independent association with SBA (aOR = 1.20, 95% CI [1.09, 1.31], p < .001). Higher parity was independently associated with lower odds of SBA; compared to women with one child, those with 6 or more children had less than half the odds (aOR = 0.48, 95% CI [0.40, 0.57], p < .001). The model demonstrated strong explanatory power (McFadden pseudo R-squared = 0.40), Multicollinearity was assessed using variance inflation factors (VIF).

All VIF values were below 6, with the highest values observed among age group dummy variables (maximum VIF = 5.52), consistent with expected correlations between categories derived from a single continuous variable. No meaningful multicollinearity was detected.

### 3.4 Confounding Assessment

Crude odds ratios were substantially larger than adjusted estimates for all predictors (Table 4). The attenuation was most extreme for higher education (crude OR = 64.04 vs. aOR = 7.01; 89.0% reduction) and richest wealth (crude OR = 48.76 vs. aOR = 6.27; 87.1% reduction), reflecting extensive confounding by correlated socioeconomic variables. Urban residence showed similarly large confounding (73.7% reduction), indicating that much of the urban-rural difference in SBA is explained by the concentration of educated and wealthier women in urban areas. ANC utilization showed the least confounding (56.3% attenuation), consistent with a more direct causal pathway between ANC contact and skilled delivery.

**Table 4:** Crude versus adjusted odds ratios: magnitude of confounding.

| Variable | Crude OR | Adjusted OR | % Change | Interpretation |
| --- | --- | --- | --- | --- |
| Higher education | 64.04 | 7.01 | -89.0% | Massively confounded by wealth and residence |
| Richest wealth | 48.76 | 6.27 | -87.1% | Highly confounded by education and urban residence |
| Secondary education | 13.02 | 2.81 | -78.4% | Substantial confounding |
| Urban residence | 4.55 | 1.20 | -73.7% | Largely explained by wealth and education |
| ANC 4 or more visits | 8.69 | 3.80 | -56.3% | Least confounded; most direct pathway |

## 4. Discussion

This nationally representative analysis of 21,465 Nigerian women identified maternal education, household wealth, ANC utilization, geopolitical region, parity, and maternal age as independent determinants of SBA after mutual adjustment. The overall SBA prevalence of 44.9% in 2018 confirms persistent gaps in Nigeria’s progress toward universal skilled delivery coverage (NPC & ICF, 2019), and the scale of regional inequality which is a 68 percentage point gap between the North West and South West, underscores the need for geographically differentiated policy responses.

Mpembeni et al. (2007) examined determinants of skilled care during delivery in Southern Tanzania and found a negative independent association between parity and skilled birth attendance utilisation, consistent with the parity findings observed in this analysis.

The finding that higher education was the strongest independent predictor of SBA (aOR = 7.01) is consistent with a substantial body of literature from sub-Saharan Africa linking maternal education to health service utilization through health literacy, decision-making autonomy, and health system familiarity (Adewuyi et al., 2017; Moyer & Mustafa, 2013). However, the 89% attenuation of the crude effect on adjustment is a critical methodological observation: studies that report crude ORs of 60 or above for education are largely capturing the effects of wealth and urban residence, not education per se. This has direct implications for how the evidence base is interpreted by policymakers. The adjusted estimate of 7.01 is still a large and policy-relevant effect, but it more accurately reflects the independent contribution of education after disentangling correlated socioeconomic factors.

The association between ANC utilization and SBA (aOR = 3.80) is among the most actionable findings of this analysis. Women who attended at least 4 antenatal care visits had nearly four times the odds of delivering with skilled care compared to those with fewer visits, even after controlling for background socioeconomic factors. This is consistent with the established role of ANC as an entry point to the formal health system: ANC contact exposes women to skilled health workers, enables birth preparedness counseling, and increases familiarity with health facilities (WHO, 2016). Crucially, ANC showed the least confounding of all predictors in this analysis (56.3% attenuation), suggesting a more direct causal pathway and reinforcing its utility as an intervention target. Scaling up ANC coverage, through free services, mobile health outreach, and community health worker programs, represents the most immediately programmable policy lever available.

The persistence of a strong independent wealth effect (aOR = 6.27 for richest vs. poorest) after adjustment for education and residence indicates that economic barriers to skilled delivery care operate through mechanisms not fully captured by education or geography alone. Direct costs of facility delivery, indirect transportation costs, and opportunity costs all disproportionately affect poor households. Conditional cash transfer programs and targeted facility delivery subsidies have shown promise in similar contexts and warrant consideration in the Nigerian policy environment.

The pronounced regional gradient, with women in the South East and South West having over six times the odds of SBA compared to the North West, even after adjustment, reflects deeply structural differences in health infrastructure density, health worker availability, and socio-cultural norms across Nigeria’s geopolitical zones. After adjustment, these regional effects are not fully explained by education, wealth, or residence, pointing to additional supply-side and community-level factors that require geographically targeted strategies. Prioritizing health systems investment in the North West and North East, where both supply and demand constraints are most severe, is a policy priority that the data support unambiguously.

The inverse association between parity and SBA (aOR = 0.48 for 6 or more children vs. 1 child) aligns with evidence on the inverse relationship between parity and skilled care utilisation in sub-Saharan African settings: higher-parity women may be less likely to seek skilled care due to prior experience with uncomplicated deliveries, reduced perceived risk, or competing household demands (Moyer & Mustafa, 2013; Mpembeni et al., 2007). Health communication strategies that address risk normalization in high-parity women may be warranted. The positive association between older maternal age and SBA, after adjustment, may reflect accumulated experience navigating the health system or the greater autonomy of older women in healthcare decision-making.

### 4.1 Strengths and Limitations

Key strengths of this study include the use of a large, nationally representative dataset with a robust sampling design, rigorous multivariable adjustment, and explicit quantification of confounding which is an analytic step that is frequently omitted in similar studies. The application of sampling weights ensures that estimates are representative of the Nigerian population of women with recent births.

Several limitations should be noted. First, the cross-sectional design precludes causal inference; the associations observed cannot confirm directionality, and reverse causation cannot be excluded for some pathways. Second, SBA was ascertained through self-report, introducing potential recall bias for births up to five years prior, and social desirability bias may lead to overreporting of skilled attendance. Third, important potential confounders, including facility availability and distance, cultural and religious norms, and women’s decision-making autonomy, were not available in the dataset and represent sources of unmeasured confounding. Fourth, the complete case exclusion of 327 women with missing ANC data assumes missingness completely at random, which was not formally tested. Finally, the data are from 2018, and the landscape of maternal care in Nigeria may have changed, particularly given health system disruptions during the COVID-19 pandemic.

## 5. Conclusions

Maternal education, household wealth, antenatal care utilization, and geopolitical region are the dominant independent determinants of skilled birth attendance in Nigeria. The substantial confounding of crude associations, particularly for education and wealth, has important implications for how evidence on SBA determinants should be interpreted and cited in policy contexts. ANC utilization stands out as the most actionable determinant given its modifiable nature and relatively direct pathway to skilled delivery. Urgent, geographically targeted investment in ANC coverage, demand-side financing mechanisms, and girls’ education are needed to close Nigeria’s SBA gap and prevent the tens of thousands of avoidable maternal deaths that occur annually.

## Data Availability

The data used in this study are publicly available from the DHS Program upon free registration. The 2018 Nigeria Demographic and Health Survey individual recode dataset (NGIR7BFL.DTA) can be accessed at: https://dhsprogram.com/data/dataset/Nigeria_Standard-DHS_2018.cfm. The analysis code is available at: https://github.com/Ugoeze-Lucy/-Determinants-of-Skilled-Birth-Attendance-in-Nigeria-A-Population-Based-Analysis-Using-DHS-2018-Data

https://github.com/Ugoeze-Lucy/-Determinants-of-Skilled-Birth-Attendance-in-Nigeria-A-Population-Based-Analysis-Using-DHS-2018-Data

